# A Virtual Skills Centre on Behaviour Management in Chronic Disease for Lay People in Low- and Middle-Income Countries (LMICs)

**DOI:** 10.1101/2020.05.10.20094060

**Authors:** Iliatha Papachristou Nadal, Nivethitha Ganapathiram, Thomas Harrison, R Deepa, Maithili Karthik, Poppy Mallinson, Chaisiri Angkurawaranon, Moffat Nyirenda, Giridhar Babu, Sanjay Kinra

**Affiliations:** King’s College London, UK; London School of Hygiene and Tropical Medicine, London, UK; Public Health Foundation of India, Bengaluru, India; Chiang Mai University, Chiang Mai, Thailand; MRC/UVRO Uganda Research Unit, Entebbe, Uganda

## Abstract

**Introduction:** Diabetes complications can be reduced and/or prevented by providing skills in behavioural management to lay people e.g. volunteers, family and friends. With a critical shortage of trained staff in lower- and middle-income countries (LMICs) to deliver these, lay people are a solution to fill this gap. Current programmes in LMICs are not suitable for lay people as they are (i) too complex i.e. for high education professionals and (ii) not easily accessible. We aim to address these two barriers through both a face-to-face and online virtual skills centre (VSC).

**Objectives:** This paper describes the development of an online VSC that will provide access to those caring for people with diabetes within a community setting; examined the suitability and usability of the VSC using qualitative methods.

**Methods:** Drawing on the DoTTI framework for developing an online program, the VSC was i) designed and developed using psychological techniques and ii) tested for early iteration using participatory workshops.

**Results:** A total of 23 participants from Bengaluru, India, both lay and clinicians, tested the VSC. The main themes identified were: empowerment for lay people; suitable for locals in a community setting; local services needed; user-friendly focused.

**Conclusion:** This study found that a theory and evidence-based approach as the basis for an online behavioural management skills programme was acceptable to users. A substantive pilot will be conducted to examine whether the VSC is effective in reducing/preventing diabetes complications and how to implement into a community setting across other LMICs and chronic diseases.

## INTRODUCTION

Diabetes and its complications are a major cause of morbidity, mortality and economical loss within lower- and middle-income countries (LMICs).^1-3^ Diabetes complications can be largely prevented through interventions provided by health care professionals such as glucose monitoring and weight management.^4^ However, there is a critical shortage of health care professionals in LMICs and as a result are unable to provide adequate diabetes management.^4-5^

A solution to this loss and shortage is through self-behavioural management programmes.^6-7^ These programmes provide the patient with the education and support to self-manage their own condition.^8^ They are delivered by professionals who are trained not just in disease specific skills but in communication and psychological skills. These programmes have shown to be effective in high income countries but in LMICs there is a lack of skilled professionals and therefore patients do not receive this form of intervention.^9-11^ As there is more informal care than formal care available, training lay people with the skills to delivery these programmes will reach more people with diabetes and hence improve their self-management.^12-13^

Lay people (i.e. community health workers, volunteers, family, friends and neighbours) are a promising resource as they understand the ethnic and local culture, live within the community and/or have a joint family structure and hence have the availability to provide these self-management programmes at a relatively low cost.^12-13^ However, current programmes in LMICs are not suitable for lay people as they are (1) too complex i.e. designed for high education professionals and (2) they are not easily accessible for lay people e.g. costly to attend or have limited free time.^14-16^ Our goal is to make a programme to address these two barriers. This will be through both a face-to-face and online virtual skills centre.

This paper describes the development of an online virtual skills centre (VSC) that will provide access to those caring for people with diabetes within a community setting and examined the suitability and usability of the VSC using qualitative methods. Key principles of the skills centre will be scalable and transferable across different chronic diseases as well as across low- and middle-income countries (LMICs).

## STUDY DESIGN

### Design and Development of Virtual Skills Centre

The DoTTI framework was used to develop and assess the virtual skills centre (VSC).^17^ This is a framework for developing web-based information tools in medicine. This framework is based on principles for developing and evaluating complex interventions,^18^ which use a phased approach, as well as drawing on the Agile methodology of software development.^19^ The DoTTI framework involves four stages from which it has taken its name: (1) design and development, (2) testing early iterations, (3) testing for effectiveness and (4) integration and implementation. This study draws on Phase 1 of the DoTTI framework. The concept and content of the intervention incorporate psychological and behavioural change techniques such as Cognitive Behavioural Therapy (CBT) and Motivational Interviewing (MI).^20-21^ These techniques aid communication skills as well as provide techniques to help with condition-specific self-management.

The development team consisted of two physicians, a health psychologist and behavioural intervention specialist from the London School of Hygiene and Tropical Medicine (LSHTM). In addition, there were two digital developers as well as public health academics from the Indian Institute of Public health (IIPH), Bengaluru, India.

The VSC is in the form of a website that can be accessed on a computer, android tablet and mobile phone. The goal of the VSC will be delivered in its final iteration as a virtual assistant application or chatbot aimed at encouraging conversational engagement among lay people in order to: a) assist on assessing psychological and behavioural changes of the person with diabetes, b) guide them through the journey of behavioural change support with personalised information, and c) improve quality and skills of those caring for people with diabetes.

**Table 1.** Components of the Virtual Skills Centre.

|  |  |
| --- | --- |
| **i** | Disease specific management skills on diabetes covering the nine care processes<sup>31</sup> adapted for culturally specific content. |
| **ii** | Communication and psychological management skills in chronic disease. |
| **iii** | An accredited course in which those completing the course will be awarded a completion certificate. |
| **iv** | The online training programme provided to participants in hard copy. |
| **v** | An online forum for peer support. |

The study received ethics approval by the London School of Hygiene and Tropical Medicine (LSHTM) ethics committee (09/11/2019) (Ref: 17755/RR/16358) and the Indian Institute of Public health (IIPH) (06/11/2019) (Ref: TRCIEC/172/2019).

### Development of an online skills centre on behavioural management

The aim of the skills centre is to provide lay people with psychological and behavioural techniques to support people managing their diabetes. This framework primarily seeks to improve the conversational element or discussions between patients and their carers in order to enhance patient self-management. The content for the online programme was developed by drawing upon evidence-based psychological techniques; focusing on MI and CBT. For the initial phase, a paper-based sample-training course was developed so as to be reviewed and evaluated with stakeholders in participatory workshops. The sample training course included four modules, are described in Table 2. One of the key requirements of the training were to ensure that the language was kept simple and clear in order to remove any assumptions of pre-existing education levels. The skills centre was translated into the local language, Kannada.

**Table 2:** Sample modules and content.

| Modules | Description of content |
| --- | --- |
| **Module 1: Introducing psychology of chronic illness:** | Understanding why it is essential to know how people may feel having diabetes and ways of managing their illness |
| **Module 2: Active listening (Part 1)** | Learning skills to be a better communicator through listening and responding to the thoughts of people with diabetes. |
| **Module 3: Open questions (Part 2):** | Learning how and when to ask the right questions, so the person with diabetes speaks in more detail about how they feel. |
| **Module 4: Positive feedback, reflections and summaries (Part 3):** | Further engaging through conversation and summarising, so both the person with diabetes and carer understanding each other. |

### Development the digital components

The website presents three key resources in both English and the local Indian language of Kannada: firstly, disease specific content about the nature, prognosis and management of diabetes; secondly, on psychological techniques for ‘patient-carer conversation’, including a Kannada voiceover and a printable version of the content to make it more widely accessible; and thirdly, an interactive online forum for lay people to seek further information and advice from each other. The website development took place in these three tranches: the development of the technology; disease specific content; and the psychological skills training. Table 3 provides a summary of the disease specific content that is deployed on the website, which followed the NICE guidelines for diabetes care processes.^22^ In addition, a key aspect of the site was to develop relevant regionalised content that would provide lay people with greater knowledge of local events that would be useful to them.

**Table 3:** Summary of disease specific content.

| Tab | Description |
| --- | --- |
| **What is Diabetes?** | Different types of diabetes and aetiology. |
| **How can I prevent diabetes complications?** | The risk factors for diabetes and information on lifestyle changes to prevent diabetes. |
| **How can I help manage diabetes?** | Different treatment options available: lifestyle changes, medication, and insulin therapy.<br>Practical information on living with the condition, such as taking blood glucose measurements. |
| **Spotting the signs: What are the complications?** | Vital end-organ complications of diabetes modelled around the NICE 9 key care processes and why these complications occur, how to spot early signs and reduce risk. |

### Initial Testing

Once the content for the VSC was developed, both disease specific information and behavioural management training manuals were circulated for review by the wider multi-disciplinary research team. Feedback regarding the perceived relevance and the clarity of the content was collected, as well as the feasibility of integrating this Web-based tool into the community care pathway. This was primarily done in order to incorporate a broad set of perspectives and expertise from their respective disciplines.

The feedback from the wider team was the need for inclusion of links to further information from local resource centres, both online and in the community. The website had to be easily localisable, not just with local language but with local content and advice.

**Figure 1:**
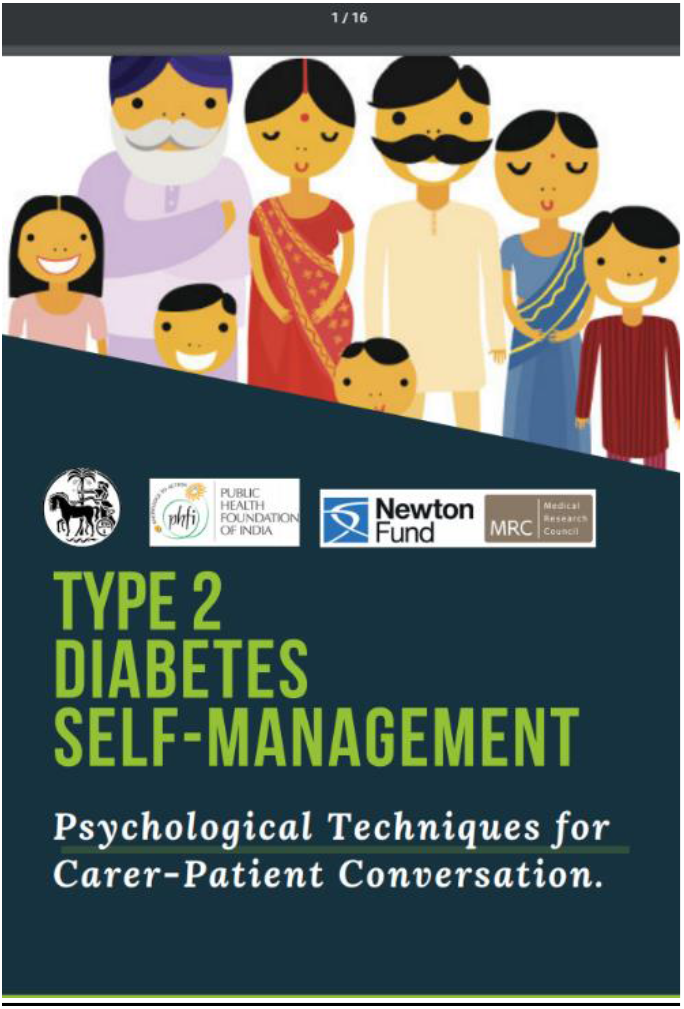
Screenshot of the front cover to the skills programme.

**Figure 2:**
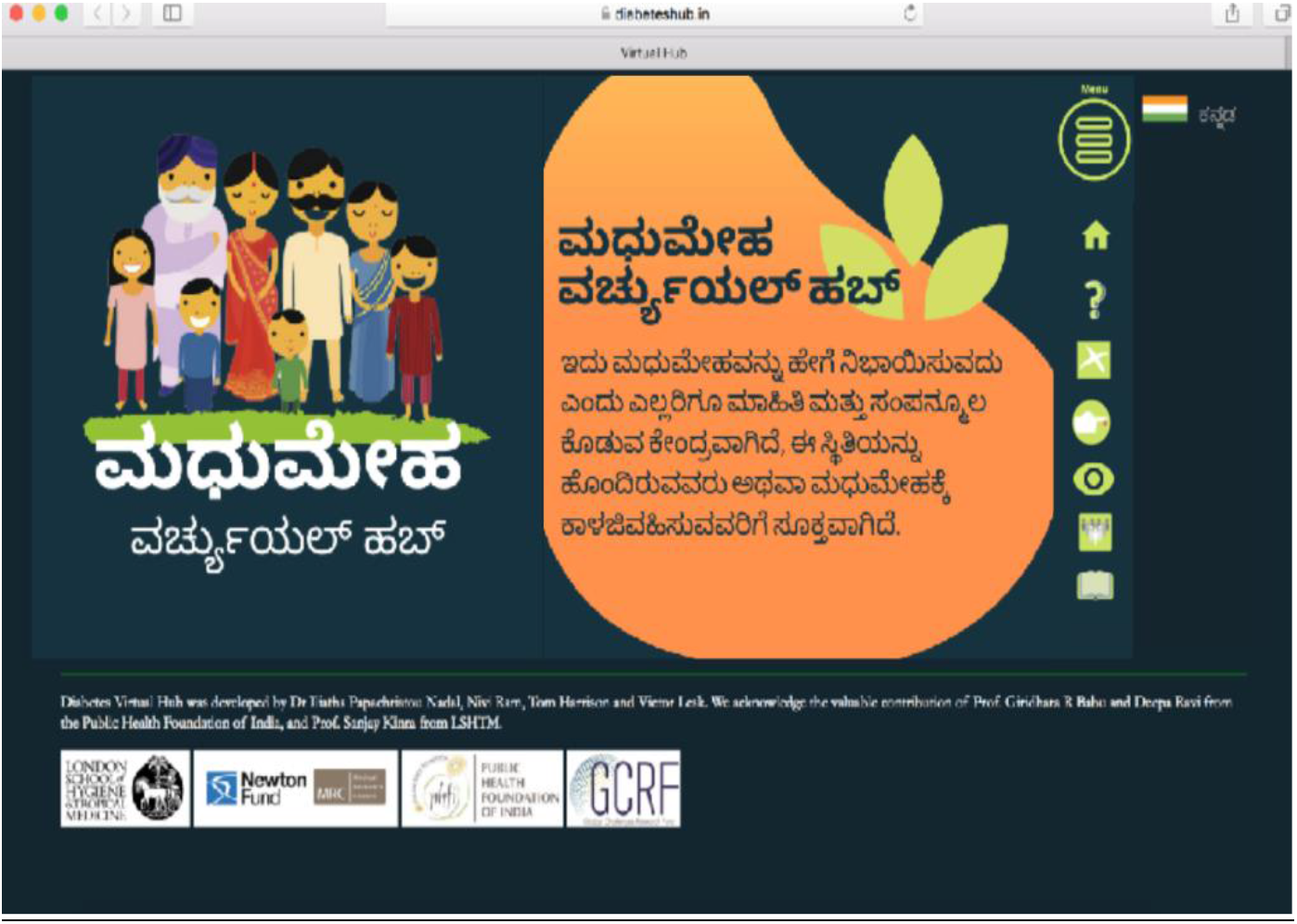
Screenshot of the website translated in Kannada.

## DATA COLLECTION

The VSC was evaluated in the form of stakeholder participatory workshops, which were used to evaluate the content and usability of the intervention while the centre was being developed. Participatory workshops create an open dialogue with stakeholders and gain as much information on their views on the content and usability of the VSC.^23^

The components of the VSC that were made available for testing were: disease specific content; a sample of the behavioural management content that was also available in audio; and a hard copy training manual.

The workshops consisted of people with diabetes and other lay people (i.e. family members; health care volunteers and workers). They were invited by the IIPH team to take part in a one-off workshop. In addition, health care professionals from the local hospitals were invited for a separate workshop.

A total of 9 health care professionals (6 medical practitioners and 3 nurses) participated in one workshop, and 14 lay people participated in another workshop, eight of which were people with diabetes (Table 4 & 5: participant demographics). The IIPH facilitated the workshops with the LSHTM overseeing the coordination and workshop topic guide. All participants were asked to navigate the website and read through the training manual. Android tablets were provided at the workshops so participants could navigate the VSC. Specifically, the two groups were asked to a) assess the content and presentation of the website b) test and assess the usability of the website c) assess the content of the sample training modules. Within each group, they were asked to discuss their thoughts on what worked and what did not work.

The workshop was facilitated using an open-structured interview schedule with three-section to assess the website and training manual. The moderators (DM and GB) guided the discussions with a topic guide (Table 6), overseen by SK. These questions were developed and refined by the authors IPN, DM, NM.

**Table 4.** Participant Demographics: Healthcare Professionals (Workshop 1)

| Healthcare Professional |  |  |
| --- | --- | --- |
| Gender | Role | Experience |
| Male | Geriatric Physician | 17 Years |
| Male | Physician | 15 Years |
| Female | OBG Physician | 37 Years |
| Female | Medicine Physician | 20 Years |
| Female | Orthopaedic Nurse | 7 Years |
| Female | Paediatric Nurse | 10 Years |
| Female | OBG Nurse | 22 Years |
| Female | OBG Nurse | 23 Years |
| Female | OBG Nurse | 21 Years |

**Table 5.** Participant Demographics: Lay People (Workshop 2)

| Family Members |  |  |  |  |
| --- | --- | --- | --- | --- |
| Gender | Age | Education | Profession | Diabetic Status |
| Male | 59 | High School | Security | 5 years |
| Male | 38 | High School | Tailor | Non-Diabetic |
| Female | 45 | No Formal Education | Maid | Non-Diabetic |
| Female | 23 | Middle School | Homemaker | Gestational Diabetes |
| Female | 43 | No Formal Education | Vendor | Non-Diabetic |
| Female | 37 | Primary School | Labourer | Non-Diabetic |
| Patients |  |  |  |  |
| Gender, Relation to family | Age | Education | Profession | Diabetic Status |
| Male, husband | 50 | Primary School | Labourer | 10 Years |
| Male, husband | 28 | Middle School | Painter | 10 Years |
| Male, husband | 50 | No Formal Education | Labourer | 4 Years |
| Female, wife | 54 | Primary School | Homemaker | 10 Years |
| Female, wife | 28 | Primary School | Homemaker | 2 years |
| Female, mother-in-law | 67 | No Formal Education | Homemaker | 10 Years |
| Lay Worker |  |  |  |  |
| Gender | Role |  | Experience |  |
| Female | Asha Worker |  | 10 Years |  |
| Female | Asha Worker |  | 8 Years |  |

**Table 6.** Topic Guide for Participatory Workshops.

|  |
| --- |
| **Website** |
| Do you think the website is useful, and why? |
| What changes/improvements would you like made to the website? |
| What more would you like to see on the website? |
| **Training Programme** |
| What do you think of the training programme? |
| What changes/improvements would you like made to the training programme? |
| What more would you add to the training programme? |

Feedback was collected through handwritten surveys from the health care professionals and oral feedback from patients, family carers and health care volunteers where facilitators actively took field notes during the session. Audio from the workshops was also collected, and flip-chart aids were used and collected to allow for post-analysis.

## DATA ANALYSIS

All data was transcribed and translated into English using a Microsoft Excel spreadsheet. The First author (IPN) used thematic analysis to code and analyse the data^24^. A second researcher (DM) then checked the codes against the data to ensure credibility and trustworthiness. This assessed how well the categories covered the data, and was discussed, defined and refined between the two authors. An iterative approach was used to develop themes that accurately reflect the data.

## RESULTS

There was a generally positive consensus from stakeholders across both workshops; with a perception that the VSC would be useful and is a much-needed resource. From the collected feedback, the main themes identified were: empowerment for lay people; suitable for local people in a community setting; local services needed; user-friendly focused.

### Empowerment for lay people

The disease specific information on the website was described by lay people as being beneficial while the health care professionals acknowledged the overall value of empowering ‘patients and their caregivers’ to self-manage. Both groups highlighted that the information on the website can address diabetes issues at any stage of the lay person’s journey:

*‘The website is very useful, as it helps a layperson to understand the importance of managing diabetes, preventing complications or even preventing the disease itself*’ (Medical Practitioner)

*“This is good as it is not just medicine, it is more for us, if we are empowered then our diabetes will be cured by 50%, the remaining 50% is medical care.”* (person with diabetes, Male)

The training modules were viewed as empowering for lay people as they felt it gave them more confidence and awareness to help the people they care for:

*“I learnt how to be more sensitive towards the patient and I feel confident to use these techniques” (Asha Worker)*

### Suitable for local people in a community setting

All stakeholders felt the skills programme is a valuable tool for lay people and that there are many people in need that will benefit from the training. In particular, having the training modules plus audio ein the local language was considered highly valuable.

*“This is ideal for health care workers and people that have a passion to improve the health of the community”* (Medical Practitioner)

*“It is useful that the local language has been introduced so that local people understand. Also, audio for the training module is a good idea.”* (Nurse)

Those caring for people with diabetes reflected on how these training skills made an impact on them: “*I feel better equipped to approach patients in the community, and I feel like I will learn better ways to interact and empathise through the new skills I’ve learnt”* (Healthcare worker).

### Local services needed

The stakeholders identified areas of improvements for the content of the website. Health care professionals mentioned that including information on local services (e.g. health and wellness centres) would increase the usage and help lay people navigate existing services. The patients thought it would be useful to have details of doctors and clinics in their community for them to contact.

*“It would be useful to have a list of professionals we can contact for specific advice or maybe there are centres we can call and visit for support”* (family caregiver)

### User-friendly focused

Feedback was received on the usability and format of the VSC and what improvements should be made. All stakeholders wanted the structure of the website to be more user-friendly and to include more visual graphic representation. The training manual specifically needed simplifying further for all lay people to understand. *“I think it needs simple language and pictures of diabetic complications and representation of exercise and dietary changes the patient should make.”* (Asha worker).

## DISCUSSION

This paper describes the development of a virtual skills centre designed for lay people with the aim to enhance diabetes behavioural management. From previous research, managing behavioural risk factors, such as poor diet and physical inactivity, can reduce and even prevent diabetes complications.^7^ Therefore, diabetes self-management education is an important pillar of diabetes care. This can be applied by enhancing lay peoples’ skills in behavioural change, psychological as well as communication techniques via face-to-face and digital platforms.^16-22^ As negative health and economic impacts of diabetes disproportionately affect poor and marginalised groups, the use of lay people combined with emphasis on social support has potential to strengthen community cohesion. In particular, in marginalised communities, it has been shown to foster wide benefits to civil society, economic and political engagement, and social development.^11-14^

The DoTTI framework method provides a systematic step by step approach that helped to develop the ‘web-based information tool’.^17^ Disease specific information and communication skills were incorporated into the virtual skills centre using simple language, images and short reflective exercises. Therefore, making it more accessible to people with low-literacy, and marginalised groups. The feedback from the participatory workshops helped to further evaluate the content, structure and functionality of the digital platform plus training materials. The stakeholders (i.e. end-users) from these workshops found the training materials to be empowering for lay people, and suitable to be used in community settings for local people. Furthermore, the training was perceived to increase lay people’ skills in psychological and communication techniques that they felt will help people with diabetes self-manage their condition.

Both health care professionals and lay people reported that through this training, lay people will have the confidence to significantly impact the lives of people with diabetes. This is because they will have the skills to give regular advice on behavioural change and provide emotional support where needed. Therefore, this programme supports current studies that found lay people who have the skills to support diabetes patients, can improve self management behaviours, such as glucose levels, diet and exercise.^13-14^ Thus, reducing complications and adverse outcomes for people with chronic conditions

### Limitations

As the intervention was tested with a small number of participants in a workshop environment rather in the community, longer-term use and post-training results were not assessed. Although, initial technological errors were tested for, i.e. usability as well as an evaluation of the content and structure, a further study is needed to test effectiveness and usability of the technology in the ‘real world’. The benefits of using small discussions workshops however, is that insight was gained from the participants shared understandings of patients and carers diabetes needs for self-management.

### Practice implications

The virtual skills centre will be modified as per the recommendations from the results of the participatory workshops. It will be refined and ready for field-testing in the form of a pilot study with aims for it to be used across other LMICs and chronic diseases. Artificial intelligence, such as in the form of a Chatbot will be developed to include a voice-to-text function. This will not only aid personalised assessments and progress but will further reach people with low literacy level that may otherwise not have access to such resources. Furthermore, the VSC has potential to have an indirect impact, through stimulating and strengthening capacity with patients’ health care providers and community support services, i.e. in both the public and private health sector.

In conclusion, providing lay people with skills to enhance psychological and behavioural management within chronic disease through accessible online skills training has the potential to address the gap in the provision of diabetes care for lay people within LMICs.

## Data Availability

All data relevant to the study are included in the article or uploaded as supplementary information. Any further data are available upon reasonable request.

## Acknowledgments

Firstly, we would like to thank the participants who took part in the workshops. Secondly, to acknowledge the two technical developers from the team: Victor Lesk and Nikolaos Maniatis (The Cato Bot Company Ltd).

## REFERENCES

1. Xie Y, Bowe B, Mokdad AH, et al. Analysis of the Global Burden of Disease study highlights the global, regional, and national trends of chronic kidney disease epidemiology from 1990 to 2016. Kid Int 2018;94(3)567–581.

2. Ogurtsova K, da Rocha Fernandes JD, Huang Y et al. IDF Diabetes Atlas: Global estimates for the prevalence of diabetes for 2015 and 2040. Diabetes Res Clin Pract 2017;128:40–50.

3. Chan M. Global status report on noncommunicable diseases. World Heal Organ 2010

4. Alaofè H, Asaolu I, Ehiri J et al. Community Health Workers in Diabetes Prevention and Management in Developing Countries. Ann Glob Heal 2017;83(3–4):661-75.

5. Lambert SD, Bowe SJ, Livingston PM, et al. Impact of informal caregiving on older adults’ physical and mental health in low-income and middle-income countries: A cross-sectional, secondary analysis based on the WHO’s Study on global AGEing and adult health (SAGE). BMJ Open 2017;7(11):1–14.

6. Vas A, Devi ES, Vidyasagar S, et al. Effectiveness of self-management programmes in diabetes management: A systematic review. Int J Nurs Pract 2017;23(5):1–8.

7. Karakurt P, Kaşikçi MK. The effect of education given to patients with type 2 diabetes mellitus on self-care. Int J Nurs Pract 2012;18(2):170–9.

8. Haas L, Maryniuk M, Beck J, et al. National standards for diabetes self-management education and support. Diabetes Educ 2012;38:619–629.

9. Mohan V, Khunti K, Chan SP. et al. Management of Type 2 Diabetes in Developing Countries: Balancing Optimal Glycaemic Control and Outcomes with Affordability and Accessibility to Treatment. Diabetes Ther 2020;11:15–35

10. Bloom DE, Chisholm D, Llopis EJ, et al. From Burden to “Best Buys”: Reducing the Economic Impact of Non-Communicable Diseases in Low- and Middle-Income Countries. World Health Organization; World Economic Forum 2011

11. Chatterjee S, Davies MJ, Heller S, et al. Diabetes structured self-management education programmes: a narrative review and current innovations. Lancet Diab End 2018;6(2):130–42.

12. Norris SL, Chowdhury FM, Van Le K. Effectiveness of community health workers in the care of persons with diabetes. Diabet Med 2006;23(5):544–56

13. Werfalli M, Raubenheimer P, Engel M, et al. Effectiveness of community-based peer-led diabetes self-management programmes (COMP-DSMP) for improving clinical outcomes and quality of life of adults with diabetes in primary care settings in low and middle-income countries (LMIC): a systematic review and meta-analysis. BMJ Open 2015;5(7): e007635.

14. Andreae SJ, Andreae LJ, Richman JS, et al. Peer-delivered cognitive behavioral training to improve functioning in patients with diabetes: A cluster-randomized trial. Ann Fam Med. 2020;18(1):15–23.

15. Esterson YB, Carey M, Piette JD, Thomas N, Hawkins M. A systematic review of innovative diabetes care models in low-and middle-income countries (LMICs). J Health Care Poor Underserved 2014;25(1):72–93.

16. Gyawali B, Bloch J, Vaidya A, Kallestrup P. Community-based interventions for prevention of Type 2 diabetes in low- and middle-income countries: a systematic review. Health Promot Int 2019;34(6):1218–30.

17. Smits R, Bryant J, Sanson-Fisher R, et al. Tailored and integrated web-based tools for improving psychosocial outcomes of cancer patients: the DoTTI development framework. J Med Internet Res 2014;16:e76.

18. Craig P, Dieppe P, Macintyre S et al. Medical Research Council Guidance Developing and evaluating complex interventions: the new Medical Research Council guidance. BMJ 2008:337:a1655.

19. Shore JH, Warden S. The Art of Agile Development. Sebastopol, CA: O’Reilly Media, Inc; 2007

20. Hofmann SG, Asnaani A, Vonk IJ, et al. The Efficacy of Cognitive Behavioral Therapy: A Review of Meta-analyses. Cog Therapy Res 2012;36(5):427–440.

21. Miller WR. Motivational interviewing with problem drinkers. Beh Psych Ther 1984;11:147–72

22. Waddingham S. Nine processes of care for diabetes. The British Journal of Primary Care Nursing 2011;8(4):171–173.

23. Kindon S, Pain R, Kesby M., eds., 2007. Participatory action research approaches and methods: connecting people, participation and place. London: Routledge

24. Braun V, Clarke V. Using thematic analysis in psychology. Qual Research Psycho 2006;3(2): 77-101.

